# Air pollution, lung function and mortality: survival and mediation analyses in UK Biobank

**DOI:** 10.1101/2023.06.23.23291813

**Authors:** Anna L Guyatt, Yutong Samuel Cai, Dany Doiron, Martin D Tobin, Anna L Hansell

**Author notes:** Corresponding author: Professor Anna Hansell, Centre for Environmental Health and Sustainability, University of Leicester, University Road, Leicester, LE1 7RH, UK. denotes joint first authorship (Anna L Guyatt, Yutong Samuel Cai).

## Abstract

**Background:** Air pollution is associated with lower lung function, and both are associated with premature mortality and cardiovascular disease (CVD). Evidence remains scarce on the potential mediating effect of impaired lung function on the association between air pollution and mortality or CVD.

**Methods:** We used data from the UK Biobank cohort (∼200,000 individuals) with 8-year follow-up to mortality and incident CVD. Exposures to PM_10_, PM_2.5_, and NO_2_ at baseline were assessed by land-use regression modelling. Lung function [forced expiratory volume in 1 second (FEV_1_), Forced Vital Capacity (FVC)] was measured by spirometry and transformed to Global Lung Function Initiative z-scores. Adjusted Cox proportional hazards and causal proportional hazards mediation analysis models were fitted, stratified by smoking status.

**Results:** Lower FEV_1_ and FVC were associated with all-cause and CVD mortality, and incident CVD. Point estimates for the mortality outcomes were larger in ever-smokers than never-smokers (all-cause mortality Hazard Ratio (HR) per decrease FEV_1_ GLI z-score, ever smokers: 1.29 [95%CI:1.24-1.34], versus never-smokers: HR 1.16 [95%CI:1.12-1.21]). Long-term exposure to PM_2.5_ or NO_2_ was associated with incident CVD, with similar effect sizes for ever and never smokers. Mediated proportions of the air pollution—all-cause mortality estimates driven by FEV_1_ were 18% [95%CI: 2%-33%] for PM_2.5_, and 27% [95%CI: 3%-51%] for NO_2_. The estimated mediated proportions for air pollution—incident CVD were 9% [95%CI: 4%-13%] for PM_2.5_ and 16% [95%CI: 6%-25%] for NO_2_.

**Conclusions:** Lower FEV_1_ may mediate some associations between air pollution on mortality and CVD outcomes, with more pronounced effect sizes for NO_2_ than for PM_2.5_.

**Take-home message:** Adverse effects of air pollution on lower lung function (FEV_1_) potentially mediate 10-30% of the effects of PM_2.5_ or NO_2_ on mortality and incident cardiovascular disease.

## Introduction

According to the Global Exposure Mortality Model, particulate matter<2.5µm (PM_2.5_) was estimated to cause nearly nine million deaths worldwide in 2015; the majority of which were from cardiovascular disease (CVD) [1]. Ambient air pollution is also an established risk factor for respiratory health. Long-term exposure to particulate matter<10µm (PM_10_), PM_2.5_ or nitrogen dioxide (NO_2_), has been associated with impaired lung function, and prevalence and incidence of chronic obstructive pulmonary disease (COPD) [2] [3] [4].

Impaired lung function is predictive of all-cause and cardiovascular mortality in general population cohorts in the United Kingdom (UK); such associations are also seen among lifelong non-smokers, as reported in UK Biobank previously [5]. These findings were supported by Mendelian randomization studies, suggesting that confounding by smoking does not explain the relationship between lung function and mortality [6] [7]. Few studies, however, have reported the potential mediating role of impaired lung function on the associations between air pollution and mortality or CVD outcomes, in particular in subgroups with different smoking status.

We previously reported in a cross-sectional analysis that ambient air pollution was associated with lower levels of lung function in UK Biobank [2]. In this present study, we further investigate the extent to which associations between air pollution and mortality or incident CVD is driven by the effect of air pollution on forced expiratory volume in 1 s (FEV_1_). FEV_1_ is a lung function measure reflecting airflow obstruction, and a lower FEV_1_ level appears to be a stronger predictor of mortality than forced vital capacity (FVC) [8]. In the largest mediation analysis of its kind, we estimated the proportions of the air pollution— outcome associations explained by the relationship between air pollution and FEV_1_.

## Methods

### Study populations

UK Biobank is a prospective cohort study of >500,000 participants (aged 40-69 years at baseline, 2006-2010), recruited from general practices across the UK [9]. On joining the study, participants attended a clinical interview with a nurse, completed questionnaires on their health and lifestyle, and performed spirometry. UK Biobank includes linkage to electronic healthcare records, as well as ambient air pollution concentration estimates at residence (**Supplementary Table 1**). All participants provided written consent and ethical approval for the UK Biobank study overall was obtained from the North West Multi-Centre Research Ethical Committee and Patient Information Advisory Group. These specific analyses have been approved as part of UK Biobank project 648.

**Table 1.** Descriptive statistics for individuals with complete data on key exposures and outcomes, versus those in two main analysis samples for mortality and incident CVD

| Factor | Level | Complete outcome data* | Mortality analysis sample | Level | CVD analysis (mortality and incidence) sample |
| --- | --- | --- | --- | --- | --- |
| N |  | 316893 | 208998 |  | 199577 |
| Outcome | Alive | 309419 (97.6%) | 204756 (98.0%) | No CVD | 189654 (95.0%) |
|  | Died (other cause) | 6301 (2.0%) | 3594 (1.7%) | Incident CVD | 9923 (5.0%) |
|  | Died (CVD) | 1173 (0.4%) | 648 (0.3%) |  |  |
| FEV<sub>1</sub> (litres), mean (SD) |  | 2.8 (0.8) (n=316893) | 2.9 (0.8) (n=208998) |  | 2.9 (0.8) (n=199577) |
| FVC (litres), mean (SD) |  | 3.7 (1.0) (n=316893) | 3.7 (1.0) (n=208998) |  | 3.7 (1.0) (n=199577) |
| FEV<sub>1</sub>/FVC, mean (SD) |  | 0.8 (0.1) (n=316893) | 0.8 (0.1) (n=208998) |  | 0.8 (0.1) (n=199577) |
| COPD (GOLD stage) | No COPD | 272468 (86.0%) | 182930 (87.5%) | No COPD | 175326 (87.8%) |
|  | Stage 1 | 20610 (6.5%) | 12850 (6.1%) | Stage 1 | 12247 (6.1%) |
|  | Stage 2 | 20994 (6.6%) | 11818 (5.7%) | Stage 2 | 10793 (5.4%) |
|  | Stage 3 | 2608 (0.8%) | 1287 (0.6%) | Stage 3 | 1119 (0.6%) |
|  | Stage 4 | 213 (0.1%) | 113 (0.1%) | Stage 4 | 92 (<1%) |
| PM2.5, 2010, median (IQR) |  | 9.9 (9.3, 10.5) (n=316893) | 9.9 (9.2, 10.5) (n=208998) |  | 9.9 (9.2, 10.5) (n=199577) |
| PM10, 2010, median (IQR) |  | 16.0 (15.2, 17.0) (n=316893) | 16.0 (15.2, 17.0) (n=208998) |  | 16.0 (15.2, 16.9) (n=199577) |
| NO2, 2010, median (IQR) |  | 25.9 (21.1, 30.9) (n=316893) | 25.6 (21.0, 30.7) (n=208998) |  | 25.6 (21.0, 30.7) (n=199577) |
| Sex | Female | 176405 (55.7%) | 116341 (55.7%) | Female | 113452 (56.8%) |
|  | Male | 140488 (44.3%) | 92657 (44.3%) | Male | 86125 (43.2%) |
| Age (at recruitment), mean (SD) |  | 56.3 (8.1) (n=316893) | 55.9 (8.0) (n=208998) |  | 55.6 (8.0) (n=199577) |
| Age group (at recruitment) | <45 | 33520 (10.6%) | 23177 (11.1%) | <45 | 23018 (11.5%) |
|  | >=45 and <55 | 92690 (29.2%) | 64090 (30.7%) | >=45 and <55 | 62926 (31.5%) |
|  | >=55 and <65 | 133931 (42.3%) | 87247 (41.7%) | >=55 and <65 | 82801 (41.5%) |
|  | >=65 | 56752 (17.9%) | 34484 (16.5%) | >=65 | 30832 (15.4%) |
| BMI, mean (SD) |  | 27.3 (4.7) (n=316623) | 27.3 (4.7) (n=208998) |  | 27.2 (4.7) (n=199577) |
| BMI category | Normal (<25 kg/m2) | 106648 (33.7%) | 70757 (33.9%) | Normal (<25 kg/m2) | 68989 (34.6%) |
|  | Overweight (25 to 29.9 kg/m2) | 134951 (42.6%) | 88967 (42.6%) | Overweight (25 to 29.9 kg/m2) | 84822 (42.5%) |
|  | Obese (≥30 kg/m2) | 75294 (23.8%) | 49274 (23.6%) | Obese (≥30 kg/m2) | 45766 (22.9%) |
| Height (cm), mean (SD) |  | 168.4 (9.2) (n=316893) | 168.6 (9.1) (n=208998) |  | 168.5 (9.1) (n=199577) |
| Education | None | 48954 (15.4%) | 26351 (12.6%) | None | 23867 (12.0%) |
|  | OLevel/CSE/GCSE | 54517 (17.2%) | 35019 (16.8%) | OLevel/CSE/GCSE | 33634 (16.9%) |
|  | A2/AS/NVQ/HNDC/Other | 106745 (33.7%) | 71546 (34.2%) | A2/AS/NVQ/HNDC/Other | 68345 (34.2%) |
|  | Degree | 103894 (32.8%) | 76082 (36.4%) | Degree | 73731 (36.9%) |
|  | Missing | 2783 (0.9%) |  |  |  |
| Income | Less than £ 31,000 | 125953 (39.7%) | 92645 (44.3%) | Less than £ 31,000 | 86623 (43.4%) |
|  | £ 31,000 and above | 148228 (46.8%) | 116353 (55.7%) | £ 31,000 and above | 112954 (56.6%) |
|  | Missing | 42712 (13.5%) |  |  |  |
| Smoking status | Never smoker | 173669 (54.8%) | 145776 (69.7%) | Never smoker | 141055 (70.7%) |
|  | Ever smoker | 143224 (45.2%) | 63222 (30.3%) | Ever smoker | 58522 (29.3%) |
| Passive smoke exposure | None | 272715 (86.1%) | 198511 (95.0%) | None | 189756 (95.1%) |
|  | Any | 15032 (4.7%) | 10487 (5.0%) | Any | 9821 (4.9%) |
|  | Missing | 29146 (9.2%) |  |  |  |
| Asthma | Never had asthma | 280868 (88.6%) | 184892 (88.5%) | Never had asthma | 176542 (88.5%) |
|  | Ever had asthma | 35564 (11.2%) | 23953 (11.5%) | Ever had asthma | 22897 (11.5%) |
|  | Missing | 461 (0.1%) | 153 (0.1%) |  | 138 (0.1%) |
| Occupation | Non "at-risk" occupation | 311072 (98.2%) | 205730 (98.4%) | Non "at-risk" occupation | 196463 (98.4%) |
|  | "At-risk" occupation | 5821 (1.8%) | 3268 (1.6%) | "At-risk" occupation | 3114 (1.6%) |

For continuous variables, numbers with non-missing data are given in brackets after the mean or median values (e.g. 2.9 (0.8) (n=208998)); for categorical variables, numbers missing are given as a separate category. \* indicates that these are the number of participants with incomplete covariate data.

### Study outcomes

The main study outcomes in this analysis include mortality (all-cause and CVD) and incident CVD. Dates and causes of death from linked death registry data were used to define all-cause and CVD mortality. Death from CVD was defined in participants with a primary cause of death specifying an ICD-10 code from the list described in **Supplementary Table 2 and Supplementary Methods**. Incident CVD (fatal and non-fatal events) was defined using the death registry data above, and from Hospital Episode Statistics data. ICD-10 codes that defined fatal and non-fatal incident CVD events are also described in **Supplementary Table 2**. Participants with prevalent CVD events at the time of entering the study were excluded. The origin for the time axis was set to year of birth, with subjects entering analysis on the date of the baseline study visit. Censor dates were as described previously [10]: censor dates for mortality were 31 January 2016 in England and Wales, and 30 November 2015 in Scotland (and for participants whose location was unclear). For incident CVD (based on death registry data *and* hospital episode statistics), censor dates were 31 March 2015 [England], 31 October 2015 [Scotland] and 29 February 2016 [Wales]. Participants with hospital admissions from multiple nations were excluded (N=2,772), as a censor date could not be confidently ascribed.

**Table 2.** Associations between three lung function variables and mortality (all-cause and CVD, N=208,998), and all incident CVD (N=199,577), stratified by smoking status

| Outcome | Subgroup | N | N outcome | FEV<sub>1</sub> z-score |  | FVC z-score |  | FEV<sub>1</sub>/FVC z-score |  |
| --- | --- | --- | --- | --- | --- | --- | --- | --- | --- |
|  |  |  |  | HR | 95%CI | HR | 95%CI | HR | 95%CI |
| All-cause mortality | Never-smokers | 145,776 | 2,267 | 1.16 | 1.12,1.21 | 1.17 | 1.13,1.22 | 1.05 | 1.00,1.11 |
|  | Ever-smokers | 63,222 | 1,975 | 1.29 | 1.24,1.34 | 1.27 | 1.22,1.33 | 1.20 | 1.14,1.25 |
| CVD mortality | Never-smokers | 145,776 | 286 | 1.31 | 1.18,1.45 | 1.38 | 1.24,1.54 | 1.01 | 0.88,1.17 |
|  | Ever-smokers | 63,222 | 362 | 1.43 | 1.31,1.57 | 1.42 | 1.29,1.57 | 1.29 | 1.16,1.43 |
| All incident CVD | Never-smokers | 141,055 | 5,743 | 1.12 | 1.10,1.15 | 1.14 | 1.11,1.17 | 1.01 | 0.98,1.04 |
|  | Ever-smokers | 58,522 | 4,180 | 1.12 | 1.09,1.15 | 1.11 | 1.08,1.14 | 1.09 | 1.05,1.13 |

\*Hazard ratios (HR) are per GLI z-score decrease for each lung function measure. Adjustments were as described in 'Covariates' of Statistical Methods.

### Air pollution estimates

Land use regression (LUR)-based estimates of PM_10_, PM_2.5_ and NO_2_ for year 2010 were linked to the geocoded residential address of each participant in UK Biobank [11] [12]. The LUR models used AirBase routine monitoring data with geospatial variables on road network, land use, population density and altitude. Performance of the LUR model was assessed by comparing the modelled estimates to those from the UK’s Automatic Urban and Rural Network monitoring data. The model performance for the LUR-NO_2_ model was relatively good (R^2^=0.67), however, for the LUR-PM_10_ model the performance was only moderately well (R^2^=0.53) for southern part of the UK. For northern England and Scotland (i.e. >400 km from Greater London), the performance for the PM model was below 0.50, therefore these areas were excluded from the PM analysis.

### Lung function measurements

Best measures of FEV_1_, FVC, and their ratio (FEV_1_/FVC), underwent quality control as described previously [13]. Pre-bronchodilation lung function tests were performed using the Vitalograph Pneumotrac 6800 spirometer (Maids Moreton, UK) by trained staff. Further outlier exclusions were undertaken by calculating per-trait z-scores, using the Global Lung function Initiative (GLI)-2012 lung function equations, with any participant with an absolute z-score value greater than five standard deviations from the mean for any trait being excluded [14]. Lung function z-scores were used in the analysis. To aid more intuitive interpretation of hazard ratios, z-scores were reflected, so that hazard ratios are given for a 1 unit *decrease* in lung function z-score.

### Statistical analysis

We first performed survival analyses investigating: i) lung function in relation to all-cause mortality, CVD mortality, and incident CVD, and ii) the relationship between air pollution and the same outcomes. Smoking-stratified analyses, and analyses in smokers adjusting for smoking heaviness were conducted to obtain more accurate estimates in ever-smokers. Effect modification by anthropometric and sociodemographic factors (including occupation), and asthma was explored. Finally, we conducted mediation analyses to examine proportions of the air pollution—outcome associations that could potentially be explained by the relationship between air pollution and FEV_1._

Rates of all-cause and CVD mortality, and incident CVD were expressed per 10,000 person-years, stratified by age group and calendar year. Rates were calculated for 2008-2015, because although data from 2006-2016 exist, there were only a small numbers of events in 2016 (since the last date of linkage in England, where the majority of participants live, was in 2015) and relatively few participants were recruited in 2006-7.

Survival analysis was performed using Cox proportional hazards models. The origin for the time axis was set to year of birth, with subjects entering analysis on the date of the baseline study visit. Proportional hazard assumptions were tested between each lung function z-score and air pollution variable with study outcomes, by plotting –ln[−ln(survival)] versus ln(analysis time) curves, per quintile of exposure. Correlations between Schoenfeld residuals and log(time) were then assessed.

Survival analyses were stratified by “ever” versus “never” smoking status and adjusted for covariates (see below): (i) FEV_1_, FVC, and FEV_1_/FVC were explored as predictors of mortality (all-cause and CVD), and incident CVD. Hazard ratios (HR) were expressed per 1-unit decrease in GLI z-score; (ii) PM_10,_ PM_2.5_, and NO_2_ were investigated as predictors of the same outcomes as above. HRs were expressed per interquartile range (IQR) increase in each pollutant.

The proportion of the associations between air pollution and the outcomes that might be driven by FEV_1_ was investigated in mediation analyses, first in all participants, then stratified by smoking status. Analyses were only conducted for pollutants with evidence of an association with the outcomes. The estimated mediated proportion was computed at the median of FEV_1_, and the exposure effect was compared across the interquartile range. Cox regression was used to model the outcome, and linear regression to model the mediator.

Survival and mediation analyses were adjusted for covariates: sex; height (cm); body mass index [BMI] in kg/m^2^; average household income before tax (dichotomised into <£31,000, and £31,000 and above); education level (categorised into “None”; “O Level, CSE or GCSE”; “A2, AS, NVQ, HNDC or Other”; and “Degree”); and passive smoking (exposure to household smoke ≥1 hour/week). In ever-smokers, results were further adjusted for smoking heaviness, using “pack-years” (1 pack year = 20 cigarettes smoked per day, per year).

Where a lung function or air pollution measure showed evidence of association with study outcomes, evidence of effect modification was assessed, using interaction terms, and then subgroup analysis, for: male vs female sex, age ≥65 vs <65 years, obesity (≥30 kg/m^2^) vs non-obesity (<30kg/m^2^), ever- vs never-smoking status, and income (≥£31,000 vs <£31,000). Covariates were as above. Interactions were additionally run in those with asthma vs no asthma, and in those having a high-risk occupation for COPD [2] [15] vs those who did not (see **Supplementary Table 3** for occupations considered).

**Table 3.**
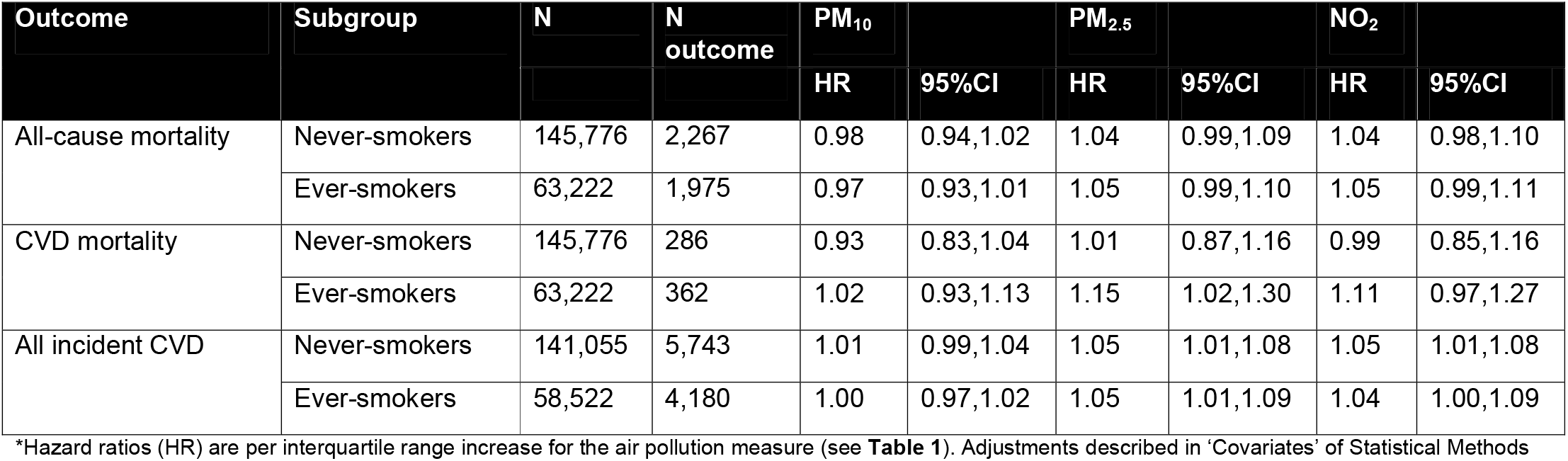
Associations between air pollution variables and mortality (all-cause and CVD, N=208,998), and all incident CVD (N=199,577), stratified by smoking status

Analysis was by complete-case analysis. Since many participants who had ever smoked had missing data for “pack-years”, two sensitivity analyses were conducted, each *omitting pack-years* from the survival analyses for lung function and air pollution. First, the pack-years variable was omitted in *ever-smokers with pack-years data*. This analysis assessed sensitivity of the results to adjusting for smoking heaviness. Next, pack-years was omitted again, but the sample of ever-smokers was expanded, this time also including *ever-smokers without pack-years data*. The latter analysis explored whether there was a selection effect when restricting to smokers with pack-years data, regardless of pack-years adjustment.

Stata 16 was used to undertake analyses, with the ‘med4way’ package used to estimate mediated proportions in a survival model framework [16].

## Results

A total of 208,998 individuals with complete covariate data (4,242 deaths, of which 648 were CVD deaths) formed the ‘mortality analysis sample’. The ‘incident CVD analysis sample’ (N=199,577, with 9,923 incident CVD events) was a subset of these individuals which excluded those with *prevalent* CVD (see study flowchart in **Figure 1**). Comparing the 316,893 individuals with incomplete covariate data to those in the mortality and incident CVD analysis samples, the latter two groups were more highly educated, had higher household incomes, and were less likely to have ever smoked (**Table 1**). Incidence rates of all-cause mortality, CVD mortality, and incident CVD were fairly static over 2008-2015, with a slight increase over time in the oldest groups (**Supplementary Figures 1-3**).

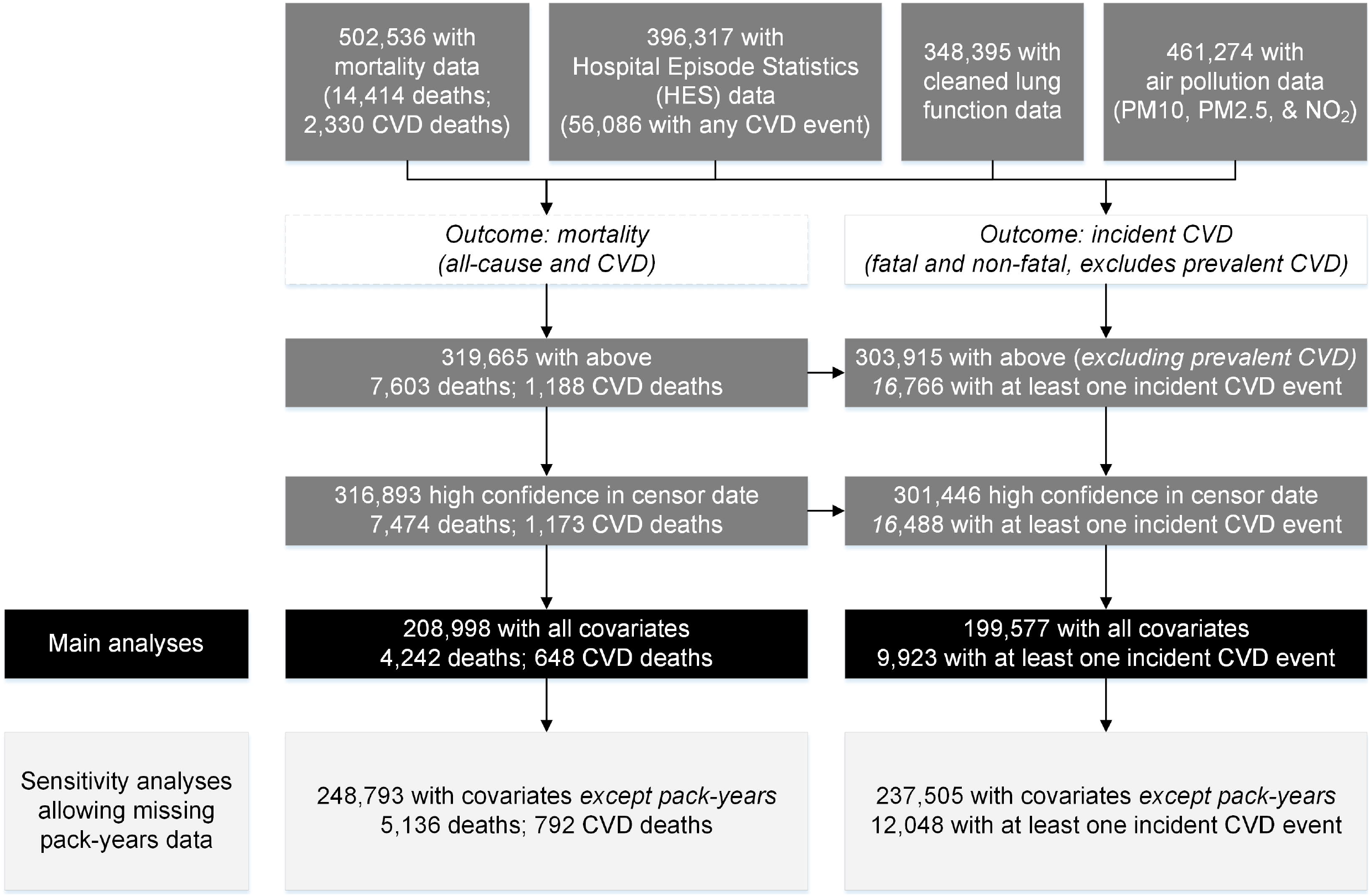

### Lung function, mortality, and incident CVD

A decrease in FEV_1_ and FVC z-scores was associated with increased all-cause mortality, CVD mortality, and incident CVD (**Table 2**). Point estimates for both mortality outcomes were larger in ever-smokers than never-smokers. For example, all-cause mortality HR per decrease FEV_1_ GLI z-score in ever smokers: 1.29 [95%CI 1.24, 1.34], versus never-smokers: HR 1.16 [95%CI 1.12, 1.21]; corresponding HRs for CVD mortality in ever smokers, FEV_1_: 1.43 [95%CI 1.31, 1.57], versus never-smokers, FEV_1_: HR 1.31 [95%CI 1.18 1.45]. Associations between FEV_1_ or FVC z-scores and incident CVD were almost identical by smoking status. For all outcomes, associations with FEV_1_/FVC z-score were found in ever-smokers only, and the relative magnitude of each association mirrored the results for FEV_1_ and FVC.

For all-cause mortality, there was evidence for a greater effect of impaired lung function in ever-smokers (HR for interaction [FEV_1_×ever-smoker]: 1.10, p=0.001), those from low-income households (HR for interaction [FEV_1_×higher-income]: 0.91, p=0.002). For incident CVD, effect sizes were slightly smaller in males (HR for interaction [FEV_1_×male sex]: 0.93, p=1.20×10^−4^) (**Supplementary Table 4**).

**Table 4.** Estimated proportions of associations between PM_2.5_ and NO_2_* and all-cause mortality and incident CVD, mediated by FEV_1_

| Outcome | Subgroup | PM<sub>2.5</sub> |  | NO<sub>2</sub> |  |
| --- | --- | --- | --- | --- | --- |
|  |  | Coefficient** | 95%CI | Coefficient** | 95%CI |
| All-cause mortality | All | 0.18 | 0.02,0.33 | 0.27 | 0.03,0.51 |
|  | Never-smokers | 0.16 | -0.05,0.38 | 0.28 | -0.12,0.67 |
|  | Ever-smokers | 0.15 | -0.05,0.35 | 0.19 | -0.04,0.43 |
| All incident CVD | All | 0.09 | 0.04,0.13 | 0.16 | 0.06,0.25 |
|  | Never-smokers | 0.11 | 0.03,0.19 | 0.20 | 0.04,0.35 |
|  | Ever-smokers | 0.06 | 0.01,0.11 | 0.10 | 0.00,0.19 |

\*Analyses not run for PM<sub>10</sub>, since there was no strong evidence of association between this pollutant and mortality or incident CVD \*\*Coefficients are estimate proportions of the effect of the pollutant on the outcome mediated via FEV<sub>1</sub>. Results were produced using the med4way Stata package, specifying the lower and upper quartiles of the air pollutants as the referent (a0) and actual (a1) levels of the exposure (a0), the median of FEV<sub>1</sub> as the level of the mediator at which the fourway decomposition is computed (m), 'cox' as the form of regression model for the outcome (yreg), and 'linear' as the form of the regression model for the mediator (mreg). Confounders (c) were fixed at female sex, income <£31,000, no qualifications, no passive smoke exposure, and at the means of height, BMI, and pack-years smoked.

### Air pollution, mortality and incident CVD

For PM_2.5_ and NO_2_, there was modest evidence of association with all-cause mortality, and associations were similar in never- and ever-smokers (**Table 3**). Point estimates for associations between PM_2.5_ or NO_2_ and CVD mortality were higher in ever-smoker, and close to the null in never-smokers. There was modest evidence of association between PM_2.5_ or NO_2_ and incident CVD, with similar associations in never- and ever-smokers. PM_10_ was not associated with either mortality or incident CVD outcomes.

There was no formal evidence of interaction between PM_2.5_ or NO_2_ and any of the covariates studied (plus asthma and occupation) in relation to study outcomes (**Supplementary Table 5**).

### Testing proportional hazard model assumptions

Proportional hazard assumptions were tested for univariate Cox regression models between quintiles of each lung function and air pollution variable with the mortality and CVD incidence outcomes (**Supplementary Figures 4-6**). There was some evidence of departure from the proportional hazards assumption for the CVD mortality analyses for FEV_1_ and FEV_1_/FVC z-scores.

### Sensitivity analyses

For lung function, removing pack-years from the model led to an increase in effect size (**Supplementary Table 6**). The point estimate was comparable when expanding the sample analysed to *all* ever-smokers (even those without pack-years data), suggesting that it was controlling for smoking heaviness (and not a selection effect in those with pack-years data), that affected the results. A similar pattern of results (albeit with less-pronounced differences) was seen for the air pollution analyses (**Supplementary Table 7, Figure 1**).

### Mediation analyses

Mediation analyses were only conducted for the associations between air pollution and all-cause mortality and incident CVD, as there was not strong evidence of an association between air pollution and CVD mortality, as seen in **Table 3**.

There was evidence that associations between PM_2.5_, or NO_2_, and all-cause mortality could be partially mediated via FEV_1_ (estimated proportion mediated: 0.18 [95%CI: 0.02, 0.33] for PM_2.5_, and 0.27 [95%CI: 0.03, 0.51] for NO_2_) (**Table 4**). For incident CVD, point estimates were larger for NO_2_ (0.16 [95%CI: 0.06, 0.25]), although confidence intervals overlapped with those of PM_2.5_ (0.09 [95%CI: 0.04, 0.13]).

## Discussion

In previous analyses of UK Biobank, we found that air pollution was associated with impaired lung function [2]. Whilst the relationship between mortality and lung function has been studied previously in lifelong non-smokers in UK Biobank [5], in the current study, we additionally studied smokers. We found lower levels of FEV_1_ and FVC were associated with all-cause mortality, CVD mortality and incident CVD regardless of smoking status, whereas associations with FEV_1_/FVC were mainly apparent in ever-smokers. We also found associations between air pollutants and mortality or incident CVD that were most prominent for NO_2_ and PM_2.5_, with no strong associations found for PM_10_. Mediation analyses suggested that FEV_1_ could mediate approximately 18% of the association between exposure to PM_2.5_ and all-cause mortality, and 27% of the association with NO_2_. Estimated mediated proportions by FEV_1_ for the associations of the same pollutants with incident CVD were smaller (9% and 16%, respectively).

Our observed associations between PM_2.5_ and NO_2_ and all-cause mortality, as well as incident CVD, were comparable to those found in the ELAPSE (Effects of Low-Level Air Pollution: A Study in Europe) project [17] [18]. The main biological mechanisms for the effects of PM or NO_2_ on morbidity and mortality proposed include promotion of oxidative stress and local pulmonary inflammation, which subsequently leads to sub-clinical systemic inflammation. Pathways involving increased oxidative stress and inflammation may drive CVD risk, via promoting atherosclerosis, endothelial dysfunction, a prothrombotic state, and alterations to the electrophysiology of the heart and blood pressure regulation [19]. Ultrafine particulates (e.g. PM_0.1_) and noxious gases (e.g. NO_2_) may also translocate/diffuse into the bloodstream and exert a direct effect on the heart and vasculature [20].

Our mediation analysis suggested that, in addition to the pathways described above, lung function impairment (measured by lower FEV_1_) may partially explain the associations between long-term exposure to ambient air pollution and mortality or incident CVD. This finding is in agreement with a previous study from the SALIA cohort of 2,527 elderly women in Germany [21]. Although the outcome assessed in the SALIA study was cardiopulmonary mortality, the confidence intervals of the proportion of the associations of NO_2_ with mortality and incident CVD mediated by FEV_1_ in our analysis are consistent with the SALIA estimate (in SALIA, 16.5% of the NO_2_—cardiopulmonary mortality association was estimated to be mediated via FEV_1_). In the SALIA study, the confidence interval of the estimated mediated proportion by FEV_1_ on PM_2.5_-cardiopulmonary mortality association overlapped the null, in contrast to our findings. Nonetheless, both studies have found that FEV_1_ mediated less of the association between PM_2.5_ and the outcomes studied, as compared to analyses of NO_2_. NO_2_ may be a marker of ultrafine particulate matter (both are generated by combustion [22]), which may cause more pulmonary inflammation and initiate an acute phase inflammatory response [23].

Our analyses demonstrated differing magnitudes of associations between lung function and mortality/CVD in ever- and never-smokers. We found that adjusting for smoking heaviness in smokers attenuated effect sizes towards those observed in never-smokers. This did not appear to be due to a selection effect of those with non-missing data for smoking heaviness. The persistent larger effect size of lung function—mortality estimates in smokers may be due to residual confounding as a result of measurement error in smoking heaviness. We also observed higher effect sizes for lung function on all-cause mortality in those with lower household incomes, and in males; a residual confounding effect by smoking heaviness may explain these findings.

To our knowledge, this is the largest mediation analysis exploring the role of FEV_1_ in driving associations between air pollution, all-cause mortality and incident CVD, and includes a more diverse population (in terms of age and sex) than studied previously [21]. We were able to stratify by smoking status, and control for smoking heaviness in analyses of smokers. The large sample size is likely to have helped detection of a potential mediating effect of FEV_1_ on PM_2.5_-mortality and CVD outcome associations; this more modest effect size for PM_2.5_ may explain why it was not detected in a previous, smaller study [21].

There are a number of limitations in this work: air pollution estimates were modelled for 2010, potentially up to four years after spirometry measures were taken (2006-2010), meaning that the mediator measurement could precede exposure measurement for some participants. However, there is evidence that air pollution measures are likely to have been relatively stable in the UK over these years [2]. Despite the larger sample size, the follow-up period was shorter than other studies, which along with the strict definition of requiring an ICD-10 as the primary cause, may have reduced the power of the CVD mortality analyses. Moreover, the analyses of CVD mortality and incident CVD may be vulnerable to bias from competing risks. Understanding causal mechanisms may require even more complex modelling of additional environmental and non-environmental confounders, such as other cardiovascular risk factors (diabetes, blood pressure) and pollutants (e.g. ultrafine particulate matter). Finally, we performed complete case analysis, since we demonstrated that covariates were unlikely to be missing at random, rendering multiple imputation difficult. Nevertheless, this will have limited the power of our analyses, and may have led to an underrepresentation of participants from more disadvantaged socioeconomic backgrounds, which may in turn introduce the possibility of collider bias [24].

In conclusion, this study suggests that the effects of PM_2.5_ or NO_2_ on lung function may account for between 10-30% of the association between these pollutants and all-cause mortality and incident CVD.

## Data Availability

This manuscript uses data from UK Biobank, available via application to the cohort https://www.ukbiobank.ac.uk/enable-your-research/apply-for-access.

https://www.ukbiobank.ac.uk/

## Acknowledgements

This study was using UK Biobank resource (project number 648). The authors would like to acknowledge Dr Mark Rutherford and Dr Lucy Teece, at Department of Health Sciences, University of Leicester, for their advice on the manuscript.

## Funding

ALG was supported by internal awards from the Wellcome Trust Institutional Strategic Support Fund (204801/Z/16/Z), and British Heart Foundation Accelerator Award (AA/18/3/34220) at the University of Leicester. MDT is supported by a Wellcome Trust Investigator Award (WT202849/Z/16/Z) and holds an National Institute of Health Research (NIHR) Senior Investigator Award.

YSC and ALH acknowledge funding from the NIHR Health Protection Research Unit in Environmental Exposures and Health, a partnership between UK Health Security Agency, the Health and Safety Executive and the University of Leicester. The views expressed are those of the authors and not necessarily those of the NHS the NIHR, UK Health Security Agency, the Health and Safety Executive or the Department of Health and Social Care.

## Author contributions

ALG was involved in all aspects of the work, including design of the analysis (with input from MDT, DD and ALH), analysis and interpretation of the data; ALG wrote the first draft of the paper, with inputs from YSC. YSC, ALH and MDT were involved in data interpretation, and all authors critically reviewed the manuscript and contributed to data interpretation and draft revision.

## Conflicts of interest

MDT has a research collaboration with GSK, and MDT and ALG have a research collaboration with Orion Pharma - all unrelated to this paper.

## Notes

### Author Declarations

Ethics committee of the North West Multi-Centre Research Ethical Committee and Patient Information Advisory Group gave ethical approval for the UK Biobank study overall (11/NW/0382), and all participants provided informed written consent for the UK Biobank study overall. The specific analyses undertaken in this paper have been approved as part of UK Biobank project 648.

