## Supplementary Methods for "Air pollution, lung function and mortality: survival and mediation analyses in UK Biobank"

ICD-10 codes used to derive CVD endpoints are described below. All codes were combined to produce one composite incident CVD endpoint. See also **Supplementary Table 2** for full code lists.

Abdominal aortic aneurysm

Codes in the category I71 (‘Aortic aneurysm and dissection’) were considered. Codes for aortic dissection, and solely thoracic aneurysms were excluded.

Atrial fibrillation and atrial flutter

Codes in the category I48 (‘Atrial fibrillation and atrial flutter’) were considered, and all were deemed relevant and retained.

Heart failure

Codes in the category I50 (‘Congestive heart failure’) were considered and all present codes included (I50.0, I50.1 and I50.9), as well as codes for hypertensive heart disease with (congestive) heart failure (I11.0), codes for hypertensive heart and renal disease with (congestive) heart failure (I13.0) and renal failure (I13.2), and pulmonary heart disease, unspecified (I27.9).

Ischaemic heart disease

Codes in the categories I20-I25 pertain to ischaemic heart disease (I20 = angina; I21 = acute myocardial infarction [MI]; I22 = subsequent MI(s) <4 weeks after index MI; I23 = complications of MI; I24 = non-specific acute IHD, of which code I24.1 is the most relevant to acute coronary syndrome [post-MI]; and I25 is chronic IHD, of which code I25.1 is most relevant to acute coronary syndromes [old MI]).

Peripheral arterial disease

Relevant categories from I70-I79 (‘Diseases of arteries, arterioles and capillaries’), pertaining to atherosclerosis, ischaemia ± infarction of the extremities were initially considered. Categories I71, I72, I77 and I78 were not deemed relevant .Relevant codes from category I70 (‘Atherosclerosis’) were those pertaining to the extremities, and other included codes were I73.1 (Buerger syndrome), I73.8 and I73.9 (related to peripheral vascular disease). Codes from I74 (‘Arterial embolism and thrombosis’) were included if they related to the extremities. I79.2 was included (‘peripheral angiopathy in diseases classed elsewhere’). Finally, relevant diabetes mellitus codes in categories E10-E14 (codes suffixed “.5”, “with peripheral circulatory problems”), were also included.

Pulmonary hypertension

Relevant codes from categories I27 (‘Other pulmonary heart diseases’) and I28 (‘Other disease of pulmonary vessels’) were initially considered. The only included codes were I27.0 (‘Primary pulmonary hypertension’) and I27.2 (‘Other secondary pulmonary hypertension’).

Stroke

Codes in categories I60-69 (cerebrovascular diseases) were initially considered. Codes in category I62 (‘Other nontraumatic intracranial haemorrhage’), including subdural and extradural haemorrhage, were excluded. Since categories I65 and I66 pertained to occlusion and stenosis of precerebral and cerebral vessels not resulting in infarction, these categories were not considered. Codes in I69 (sequelae of stroke) were used to exclude prevalent disease only, and not to define incident events.

Codes were further subdivided into ischaemic versus haemorrhagic stroke codes. Codes related to sequelae of stroke (subarachnoid or intracerebral haemorrhage, or cerebral infarction), in category I69, were used to exclude prevalent disease.

### Supplementary Figure 1

Incidence of all-cause mortality (per 10,000 person years) between 2008-2015, stratified by age group at baseline

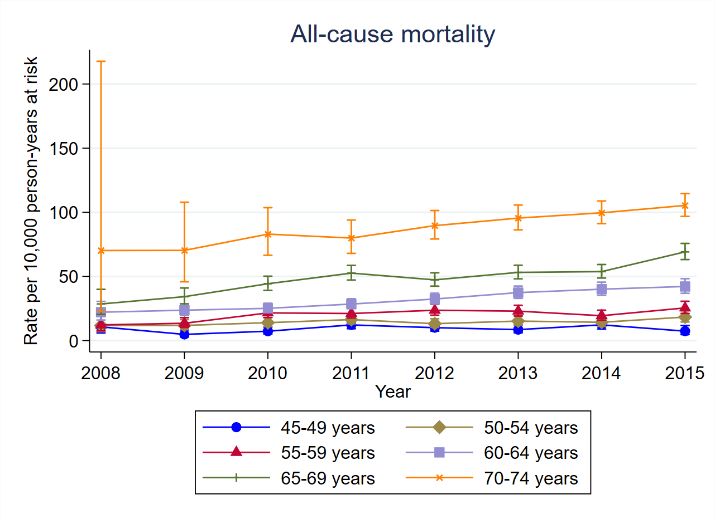

### Supplementary Figure 2

Incidence of cardiovascular mortality (per 10,000 person years) between 2008-2015, stratified by age group at baseline

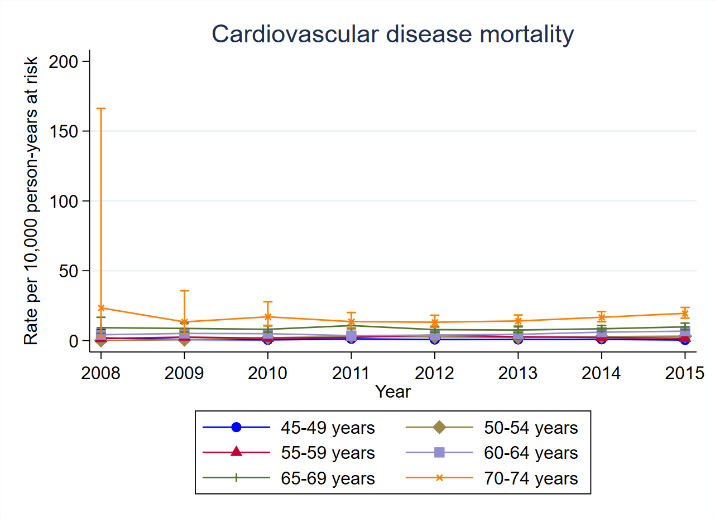

### Supplementary Figure 3

Incidence of cardiovascular events (fatal and non-fatal) (per 10,000 person years) between 2008-2015, stratified by age group at baseline

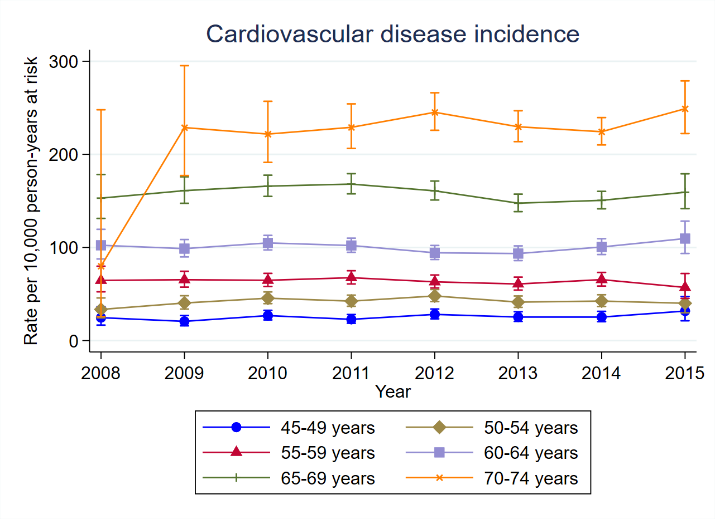

### Supplementary Figure 4

Outcome: all-cause mortality. −ln{−ln(survival)} vs ln(analysis time) curves for each quintile of exposure, for all lung function and air pollution variables. P-values are for the output of the Stata command ‘estat phtest’ (global test of correlation of Schoenfeld residuals with log(time)

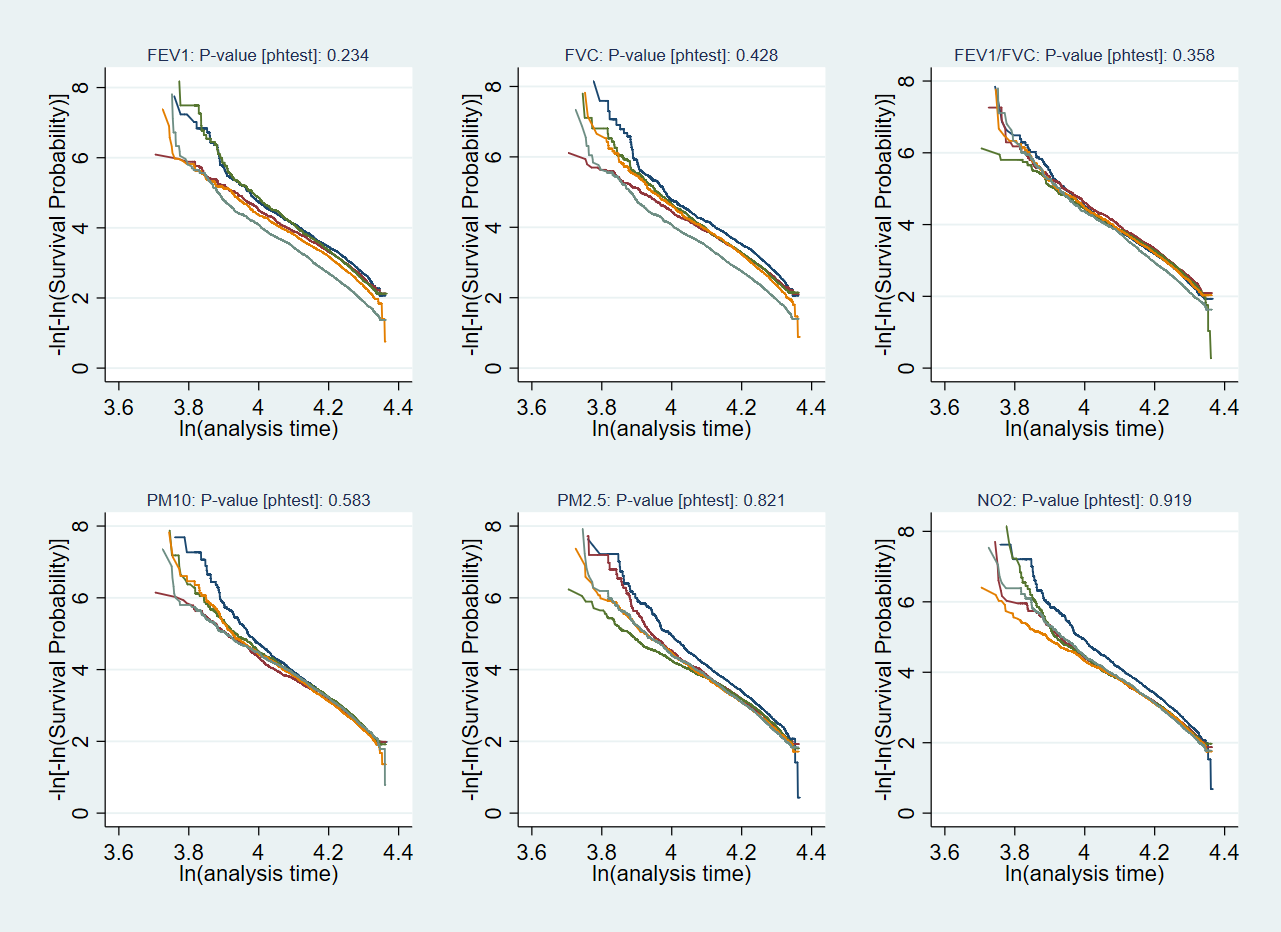

### Supplementary Figure 5

Outcome: CVD mortality. −ln{−ln(survival)} vs ln(analysis time) curves for each quintile of exposure, for all lung function and air pollution variables. P-values are for the output of the Stata command ‘estat phtest’ (global test of correlation of Schoenfeld residuals with log(time)

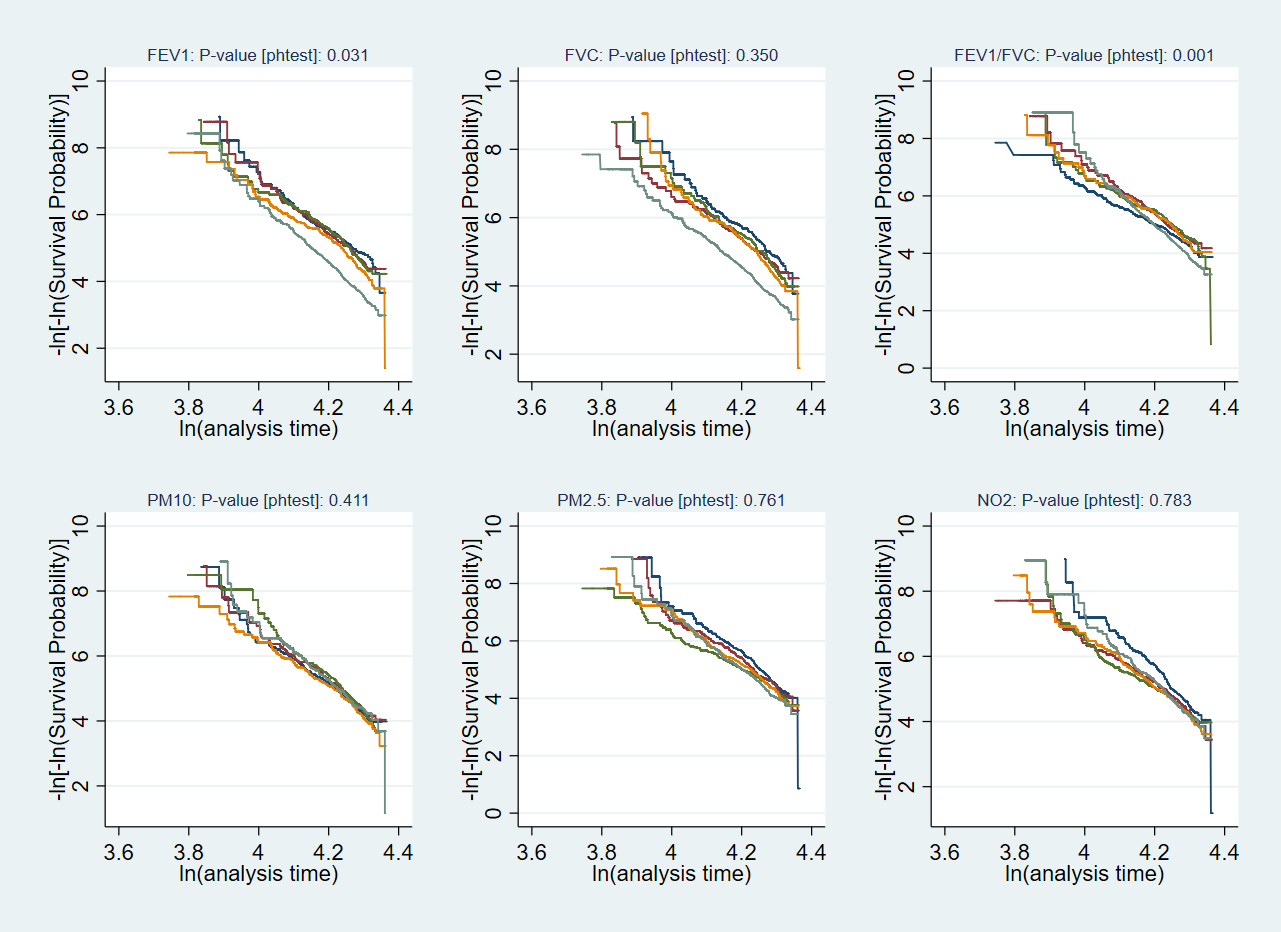

### Supplementary Figure 6

Outcome: all incident CVD. −ln{−ln(survival)} vs ln(analysis time) curves for each quintile of exposure, for all lung function and air pollution variables. P-values are for the output of the Stata command ‘estat phtest’ (global test of correlation of Schoenfeld residuals with log(time)

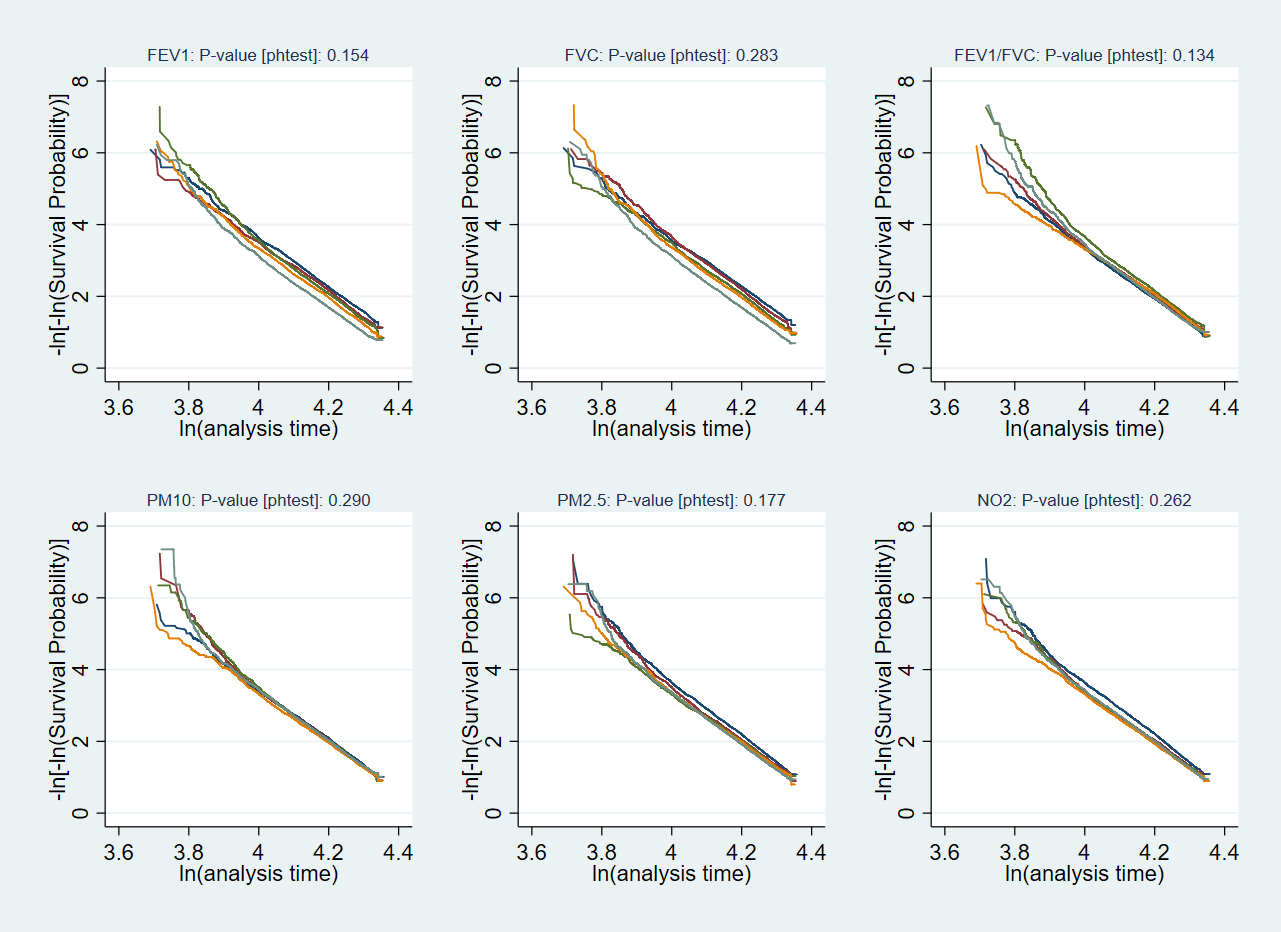
